# Atypical atrial flutter: a new methodology for critical isthmi identification?

**DOI:** 10.1101/2020.04.24.20077461

**Authors:** Pedro Adragão, Daniel Matos, Francisco Moscoso Costa, Pedro Carmo, Diogo Cavaco, Gustavo Rodrigues, João Carmo, Francisco Morgado, Miguel Mendes

**Affiliations:** Cardiology and Electrophysiology Department, Hospital de Santa Cruz, Av. Prof. Reinaldo dos Santos, 2790-134 Carnaxide, Lisbon, Portugal

**Keywords:** Atypical atrial flutter, catheter ablation, critical isthmus, high-density electroanatomic mapping, local activation times histogram

## Abstract

**Introduction:** Atypical atrial flutter is a supraventricular arrhythmia that can be treated with catheter ablation. However, the best approach is still to be defined and this strategy has suboptimal results. The Carto^®^ electroanatomical mapping (EAM) system can display a histogram of the local activation times (LAT) of the tachycardia cycle length (TCL). This study aimed to assess the ability of this new tool to identify the critical isthmus of this arrhythmia.

**Methods:** Retrospective analysis of a unicentric registry of individuals who underwent left AFL ablation during a 1-year period with Carto^®^ EAM. All patients with non-left AFL, lack of high-density EAM, less than 2000 collected points or lack of mapping in any of the left atrium walls or structures were excluded. We compared the ablation sites of arrhythmia termination to the sites of histogram valleys (LAT-Valleys), defined as areas of low-voltage (<0.3mV) with 10% or more of the TCL and less than 20% density points relative to the highest density zone. The longest LAT-Valley was designated as the primary valley, while additional valleys were named as secondary.

**Results:** A total of 9 patients (6 men, median age 75 IQR 71-76 years) were included. All patients presented with left AFL and 66% had a previous atrial fibrillation and/or flutter ablation. The median TCL and number collected points were 254 (220─290) milliseconds and 3300 (IQR 2410─3926) points, respectively. All AFL presented with at least 1 LAT-Valley in the analysed histograms, which corresponded to heterogeneous low-voltage areas (<0.3mV). All arrhythmias were effectively terminated after undergoing radiofrequency ablation in the primary LAT-Valley location.

**Conclusion:** A low-density and prolonged LAT-Valley in a heterogeneous low-voltage area compose an electrophysiologic triad that allows the identification of the AFL critical isthmus. Further studies are needed to assess the usefulness of this tool for improving catheter ablation outcomes.

## Abstract

**Introdução:** O flutter auricular (AFL) atípico é uma arritmia supraventricular que pode ser alvo de tratamento por ablação com cateter. No entanto, a melhor abordagem ainda não foi definida e esta estratégia apresenta resultados subótimos. O sistema de mapeamento eletroanatomico (EAM) Carto^®^ pode exibir um histograma dos tempos de ativação local (LAT) do ciclo do FLA. Este estudo tem como objetivo avaliar a capacidade de identificação do istmo crítico desta arritmia com esta nova ferramenta.

**Métodos:** Análise retrospetiva de um registo unicêntrico de doentes submetidos a ablação de AFL esquerdo durante um período de 1 ano e EAM com Carto^®^. Foram excluídos todos os pacientes com FLA não esquerdo, ausência de EAM de alta densidade, aquisição de menos de 2000 pontos ou falta de mapeamento em qualquer parede ou estrutura da aurícula esquerda. Os locais de ablação nos quais houve supressão das arritmias foram comparados com os vales identificados nos histogramas, definidos como zonas de baixa voltagem (<0.3mV) com 10% ou mais do ciclo taquicardia e menos de 20% da densidade de pontos EAM relativamente à zona de maior densidade. O vale do LAT mais longo foi designado como vale primário, enquanto vales adicionais foram nomeados como secundários.

**Resultados:** Um total de 9 doentes (6 homens, mediana de 75 anos IQR 71-76 anos) foram incluídos. Todos os pacientes apresentaram AFL esquerdo e 66% foram sujeitos a ablação de fibrilhação e/ou flutter auricular prévias. O ciclo mediano da taquicardia e o número mediano de pontos adquiridos foram de 254 (220─290) milissegundos e 3300 (IQR 2410─3926) pontos, respetivamente. Todos os AFL apresentaram pelo menos um vale primário nos histogramas analisados, todos correspondendo a áreas heterogéneas de baixa voltagem (<0.3mV). Todas as arritmias foram efetivamente tratadas após a ablação por radiofrequência no vale primário.

**Conclusão:** Vales prolongados e profundos no histograma do LAT em zonas de voltagem baixa e heterogénea constituem uma tríade eletrofisiológica que permite a identificação dos istmos críticos dos AFL. Mais estudos são necessários para avaliar a utilidade desta ferramenta na melhoria dos resultados da ablação por cateter.

## INTRODUCTION

Atrial flutter (AFL) is a supraventricular tachycardia that arises from both substrate and ablation scars^1,2^. While typical AFL is effectively treated with cavotricuspid isthmus ablation^3^, catheter radiofrequency (RF) ablation results are suboptimal in patients with atypical AFL^4,5^, especially when the macrorrentrant circuit involves the left atrium. Left AFL ablation strategy usually employs electroanatomic mapping (EAM) systems and entrainment mapping^6^. Another possibility is the identification and ablation of the mid-diastolic isthmus^7,8^. Nonetheless, the lack of a precise tool to identify the tachycardia critical isthmus may be one of the reasons of the technique mixed results.

## METHODS

### Patient population and study design

We conducted a retrospective analysis of consecutive patients undergoing catheter ablation for left AFL in the previous year. The patients were identified from the prospective registry for which all patients gave written informed consent.

Inclusion criteria were defined as:

- EAM with the Carto^®^ system;
- At least 1 well distributed high-density activation and voltage map during the procedure;
- More than 2000 evenly collected points;

### Electrophysiological study

All patients were anticoagulated for at least 1 month before the procedure and transesophageal echocardiography or computed tomography angiography were performed before the procedure to exclude left atrial thrombus. The procedures were performed under an activated clotting time of over 300 seconds, an octopolar steerable catheter was placed in the coronary sinus (CS), and a 20 poles catheter (Pentaray^®^, Biosense Webster Inc, California, USA) was used for high-density mapping. Transseptal puncture was performed with a Brockenbrough needle and a SL 1 sheath (Abbott Laboratories, Abbott Park, IL, EUA). The mapping was performed with Carto^®^ EAM system 6.0 version with the CONFIDENSE™ Module with high-density mapping. The activation map was created with a coronary sinus fixed reference. Wavefront algorithm was used to signal annotation. Geometry was created with fast anatomical mapping and a definition of 17. The map was collected based on the following CONFIDENSE filters settings: Tissue Proximity Either Electrode or Internal Points Filter set to 7, position and LAT stability of 4, density of 1mm and respiratory gating. RF energy was delivered with a steerable 3.5mm irrigated, contact-force catheter (Thermocool^®^, Biosense Webster Inc, California, USA). All patients underwent conventional mapping, entrainment, scar homogenization and/or lines of conduction block. The ablation was performed during AFL rhythm. In patients submitted to previous pulmonary vein isolation, residual gaps were eliminated to restore bidirectional block.

### AFL isthmus evaluation: EAM with LAT Histogram analysis

A post-hoc analysis was done with Carto^®^ version 7.0. This new model displays the local activation time sequence in a histogram of 20 bars, each one corresponding to 1/20 of the TCL. The amount of points per bar represents the segmental activated area of left atrium with the same LAT (1/20 TCL). In a previous study with other EAM system^9^, a decrease in LAT, defined as a “valley” in the LAT histogram, was related with areas of slow conduction. Based on this report, we classified a valley in the LAT histogram (LAT-Valley) as a section of the plot with 20% or less recorded points than the maximum LAT value (highest bar). To accept this section as zone of slow conduction area, it should contain at least 10% of the TCL, the substrate of this zone should be a heterogeneous scar tissue and with a local voltage in between 0.05mV to 0.3mV. These three criteria constitute an electrophysiological triad that identifies an AFL isthmus. When the LAT histogram presents more than 1 “valley”, we designate as “primary LAT-Valley” the area with the longest LAT duration, and all remaining ones are as “secondary LAT-Valleys”. We retrospectively analysed the activation and voltage maps and compared the site of AFL termination during catheter ablation with the identified LAT-Valleys. The Carto^®^ area measurements feature was used to assess the isthmus’ dimension. Normally and non-normally distributed variables were expressed as means ± standard deviation and median, respectively.

## RESULTS

Out of 22 patients, 9 fit the criteria, 3 being female, with a median age of 75 IQR 71 ─ 76 years. The clinical and arrhythmic characteristics of the 9 patients were included in this study are presented in Tables 1 and 2.

**Table 1.** Baseline patient characteristics; *including pulmonary vein isolation

| **Baseline patient characteristics** |  |
| --- | --- |
| Age, years (median, IQR) | 75 (69–76) |
| Female gender, n (%) | 3 (33%) |
| <u>Cardiovascular risk factors</u> | not applicable |
| Body mass index, n (median, IQR) | 28 (25–29) |
| Current or previous smoking, n (%) | 3 (33%) |
| Hypertension, n (%) | 8 (89%) |
| Diabetes Mellitus, n (%) | 4 (44%) |
| Dyslipidemia, n (%) | 4 (44%) |
| History of stroke, n (%) | 1 (11%) |
| History of myocardial infarction, n (%) | 0 |
| History of heart failure, n (%) | 0 |
| History of chronic kidney disease, n (%) | 0 |
| CHA<sub>2</sub>DS<sub>2</sub>-VASc Score, (median, IQR) | 3 (1–4) |
| Previous atrial fibrillation ablation, n (%) | 6 (67%) |
| Previous typical atrial flutter ablation, n (%) | 2 (22%) |
| Previous atypical atrial flutter ablation, n (%) | 1 (11%) |
| *Arrhythmia characteristics* | not applicable |
| Tachycardia cycle length, milliseconds (median, IQR) | 254 (220–290) |
| Collected electroanatomical points, n (median, IQR) | 3300 (2410–3926) |
| Primary valley duration, milliseconds (median, IQR) | 81 (51–84) |
| Valleys, n (median, IQR) | 1 (1–2) |
| Perimitral flutter, n (%) | 2 (22%) |
| Roof scar-related flutter, n (%) | 2 (22%) |
| Anterior scar-related flutter, n (%) | 1 (11%) |
| Posterior scar-related flutter, n (%) | 1 (11%) |
| Right superior pulmonary vein-related flutter, n (%) | 1 (11%) |
| Isthmus dimension, cm2 (median, IQR) | 1.97 (1.62–2.74) |
| Procedure duration, minutes (median, IQR) | 141 (105–208) |
| Radiofrequency duration*, minutes (median, IQR) | 26 (4–51) |

**Table 2:** patients’ arrhythmia characteristics

| Patient | Sex | Age (years) | AFL re-entry localization | Nr of EAM points | TCL (ms) | Number of LAT-Valleys | Primary LAT-Valley duration (ms) | Secondary LAT-Valley duration (ms) |
| --- | --- | --- | --- | --- | --- | --- | --- | --- |
| 1 | Female | 77 | Ridge | 2002 | 240 | 1 | 84 | 0 |
| 2 | Male | 74 | Perimitral | 3302 | 325 | 2 | 81 | 49 |
| 3 | Male | 75 | Perimitral | 2398 | 250 | 1 | 88 | 0 |
| 4 | Male | 48 | Ridge | 4229 | 200 | 2 | 50 | 20 |
| 5 | Male | 75 | Roof | 2720 | 210 | 2 | 84 | 42 |
| 6 | Female | 72 | Roof | 3841 | 255 | 2 | 38 | 26 |
| 7 | Male | 71 | Anterior | 3817 | 320 | 2 | 80 | 58 |
| 8 | Female | 76 | Posterior | 4011 | 260 | 1 | 52 | 0 |
| 9 | Male | 76 | LSPV | 2421 | 230 | 2 | 81 | 40 |

Six out of the 9 patients had already have a previous pulmonary vein ablation, 2 of them had a previous typical AFL ablation and 1 patient had a previous mitral AFL ablation (table 2). Three of patients had never performed a catheter ablation procedure. Perimetral (n=2), roof scar-related (n=2), anterior scar-related (n=1), posterior scar-related scar (n=1) and right superior pulmonary vein-related flutter (n=1) were identified as the mechanisms of the arrhythmias. The AFL had a median TCL of 254 (interquartile range [IQR] 220 ─ 290) milliseconds and the EAM were constituted of a median 3300 (IQR 2410 ─ 3926) points and presented a median of 2 (IQR 1 ─ 2) LAT- Valleys. The primary valleys median duration was of 81 (IQR 51─ 84) milliseconds.

All 9 patients’ AFL were effectively terminated during RF application and the arrhythmia termination zone was tagged in 3-dimensional map (Figure 1, 2 and 3). Post-hoc showed that all these areas corresponded to the primary LAT-Valley identified in the global histogram analysis. All these areas corresponded to small atrial surfaces of 1 to 2cm^2^ with heterogenous low-voltage tissue (between 0.05 and 0.3mV).

**Figure 1:**
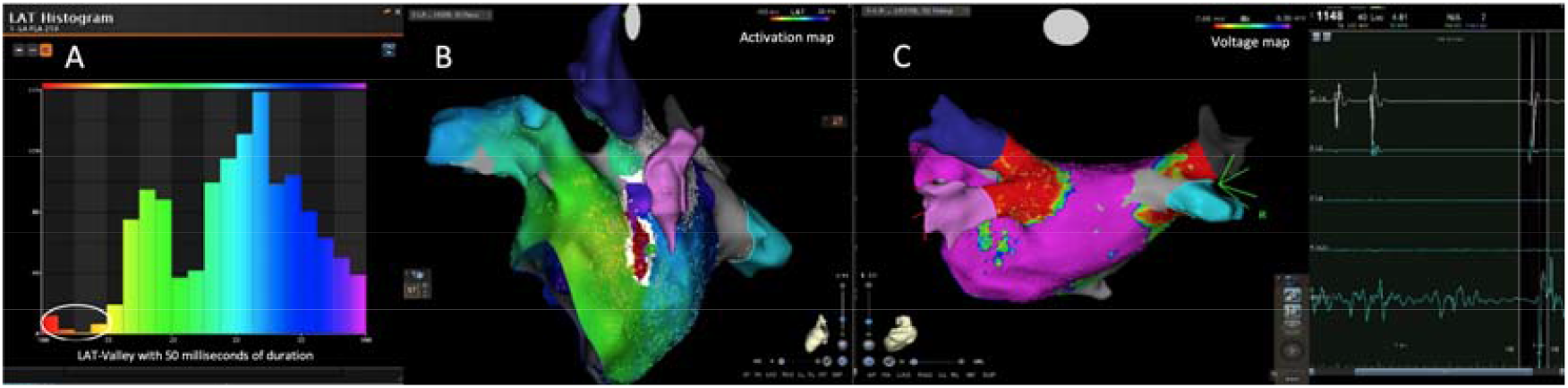
Posterior wall-related flutter, with a valley in the LAT histogram of the global activation (A) that corresponded to a delayed conduction area on the activation map (B), and a scar surrounding a channel of healthier tissue on the voltage mapping (C); the arrhythmia terminated during radiofrequency applications on this area.

**Figure 2:**
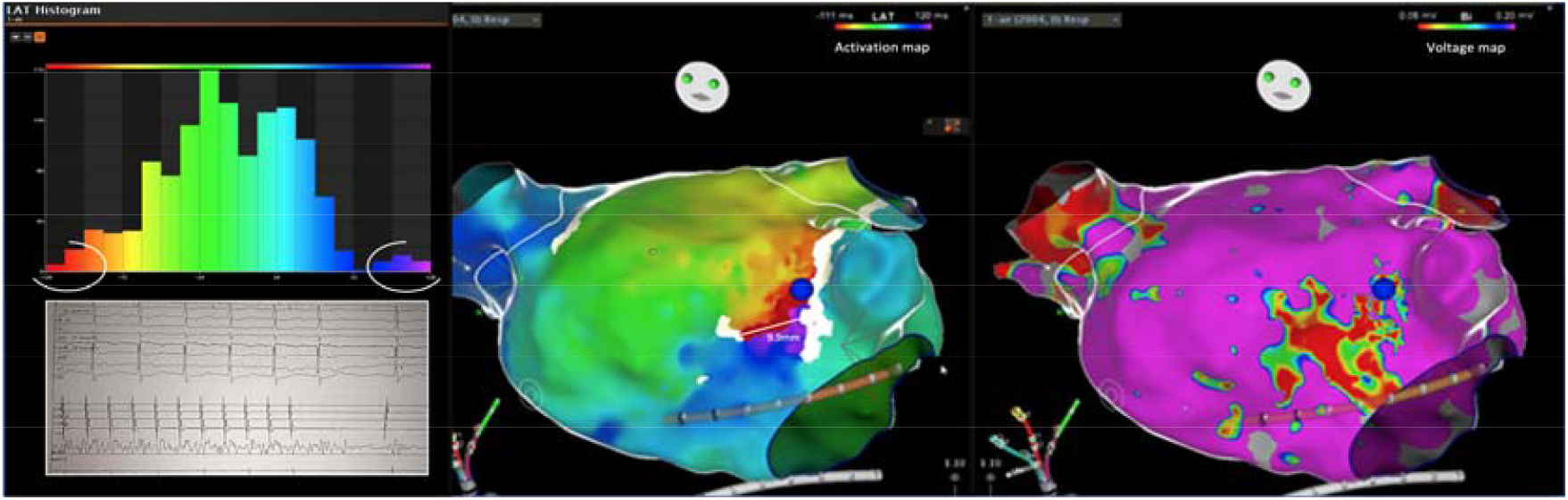
Flutter around an anterior wall scar and a corresponding LAT-Valley (blue, purple, red and orange bars); RF application stopped the AFL (blue tag).

**Figure 3:**
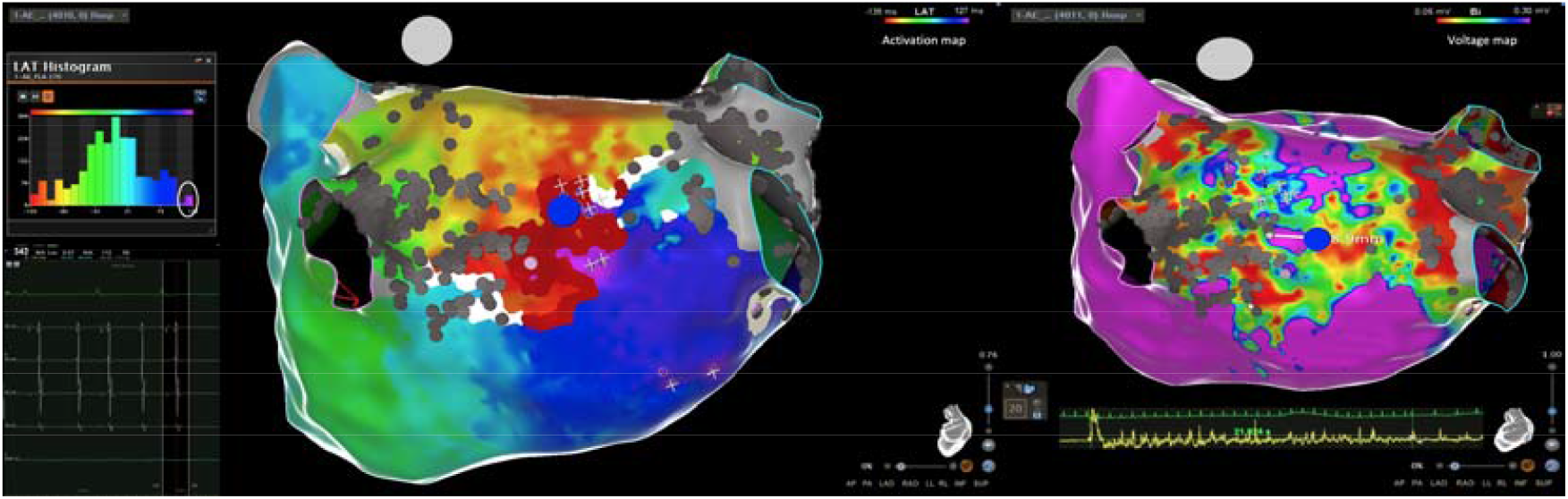
Posterior wall-related flutter, with a critical isthmus in and heterogeneous low-voltage, corresponding to a primary valley (magenta bar); the arrhythmia was stopped after 21 seconds of RF application.

## DISCUSSION

Previous approaches to atypical AFL ablation show heterogeneous catheter ablation approaches^10,11^. Most centres combine EAM with entrainment manoeuvres in order to identify critical areas of the reentrant circuit^6,11,12^. More recently, some authors suggest the identification of the mid-diastolic isthmus (MID) as a potential ablation target^7,8^. We also used conventional techniques for mapping and ablation of these patients. However, the procedures have high complexity, with extensive lesions and long duration. The results are suboptimal^6^, indicating that such techniques are not enough to identify all the critical isthmi. In our post-hoc analysis, we identified a triad that apparently recognizes a critical isthmus: (1) LAT-Valley with ≤20% the EAM points density relative to the highest density zone histogram, (2) LAT-Valley encompassing ≥10% of the TCL, (3) and a low-voltage heterogeneous area (0.05 to 0.3mV). These patterns are linked to a limited atrial surface activation and consequently to a slow conduction area (of 1 to 2cm^2^). We also showed the greater the amount of the TCL encompassed in the LAT-Valley, the highest is the probably of identifying the critical isthmus. The higher voltage in the heterogeneous scar should be the ideal ablation target. In the first seven cases the exact locations for ablation were defined in a standard fashion, whereas in the last two cases the ablation sites were selected accounting for cycle length duration and heterogenous scar (2 out of 3 triad-defined elements). In both these cases a single RF application suppressed the AFL. In a previous report^9^ with another mapping system (Rhythmia^®^), a valley in the global late activation histogram showed correlation with a critical isthmus area. However, a possible limitation could be an incorrect and insufficient point collection, that induces false positive results. In this situation, scar tissue with delayed conduction would not be present. The triad presented in this study avoids this possibility since heterogeneous low-voltage (0.05 to 0.3mV) and conduction delay (≥10% of TCL included) need to present in the LAT-Valley, increasing its specificity and accuracy. The analysis of the activation maps also showed a secondary LAT-Valley in most of the patients, a finding not previously reported. This is explained by the occurrence of more than 1 atrial area with analogous characteristics of the primary valley. Multiple slow areas are present in more complex AFL. This increased complexity is related to multiple iatrogenic scars, as seen in the voltage mapping, resulting in complex macrorrentrant circuits. In this regard, the secondary LAT-Valleys may be a potential therapeutic target if ablation of the primary valley is not sufficient. This electrophysiologic triad may be recognized and evaluated in any scar-related reentrant tachycardia. In the future, these electroanatomic isthmi may be easily identified in the 3-dimensional map, to improve and to simplify the ablation procedure.

### Study limitations

The limitations of this study should be recognized. This was a unicentric retrospective study of patients undergoing atypical AFL. Our analysis only assessed patients with effective termination of the arrhythmia, which may have induced a “survival bias” phenomenon. Also, the reduced sample of this study should be taken in consideration in the generalization of our findings. Also, we lack long-term follow up for arrhythmia relapse. A randomized study should be performed to clarify whether this tool can improve catheter ablation outcomes.

## CONCLUSIONS

A low-density and prolonged LAT-Valley in a heterogeneous low-voltage area compose an electrophysiologic triad that allows the identification of the AFL critical isthmus. Further studies are needed to assess the usefulness of this tool for improving catheter ablation outcomes.

## Data Availability

Not available for the timebeing

## ETHICAL STANDARDS

All human and animal studies have been approved by the appropriate ethics committee and have therefore been performed in accordance with the ethical standards laid down in the 1964 Declaration of Helsinki and its later amendments.

The patients signed an informed consent both for the procedure and publication of any relevant data.

